# Stochastic modelling of the effects of human-mobility restriction and viral infection characteristics on the spread of *COVID-19*

**DOI:** 10.1101/2020.07.28.20163980

**Authors:** Shiho Ando, Yuki Matsuzawa, Hiromichi Tsurui, Testuya Mizutani, Damien Hall, Yutaka Kuroda

## Abstract

After several weeks of “lockdown” as the sole answer to the *COVID*-19 pandemic, many countries are restarting their economic and social activities. However, balancing the re-opening of society against the implementation of non-pharmaceutical measures needed for minimizing interpersonal contacts requires a careful assessment of the risks of infection as a function of the confinement relaxation strategies. Here, we present a stochastic coarse grained model that examines this problem. In our model, people are allowed to move between discrete positions on a one-dimensional grid with viral infection possible when two people are collocated at the same site. Our model features three sets of adjustable parameters, which characterize (i) viral transmission, (ii) viral detection, and (iii) degree of personal mobility, and as such, it is able to provide a qualitative assessment of the potential for second-wave infection outbreaks based on the timing, extent, and pattern of the lockdown relaxation strategy. In line with general expectations, our model predicts that a full lockdown yields the best results, namely, the lowest number of total infections. A less anticipated result was that when personal mobility is increased beyond a critical level, the risk of infection rapidly reaches a constant value, which depends solely on the population density. Furthermore, according to our model, confinement alone is not effective if it is not accompanied by a detection capacity (coupled with quarantine) that surpasses 40% of the patients during their symptomatic phase. The results of our simulation also showed that keeping the virus transmission probability to less than 0.4, which can be achieved in real life by respecting social distancing or wearing masks, is as effective as imposing a mild lockdown. Finally, we note that detection and quarantine of pre-symptomatic patients, even with a probability as low as 0.2, would reduce the final numbers of infections by a factor of ten or more.

**Availability:** http://domserv.lab.tuat.ac.jp/COVID19.html (under preparation)

## Introduction

*COVID*-19 disease is an ongoing pandemic that was initially identified in Wuhan, China, in December 2019[1, 2]. Since then, until the time of writing (July 2020), over fourteen million *COVID-* 19 patients have been reported worldwide, resulting in more than **600**,**000** fatalities [3]. *COVID*-19 infection is thought to occur from close person-to-person contact [4]. This has led most countries to enforce some kind of confinement policy to reduce interpersonal contact, thereby slowing the spread of infection in the absence of a specific medical anti-*COVID*-19 treatment [5].

The causative agent of *COVID*-19 is *SARS-Cov2 (Severe Acute Respiratory Syndrome Coronavirus 2)*, a lethal member of the *Coronaviridae* family which has been a focus of much attention since 2002 [6, 7]. *SARS-CoV2* is thought to be transmitted through encounters of small droplets produced by coughing or sneezing by an infected patient and dispersed either in the air (aerosols) or on surfaces (fomites) [5]. Aerosols are either directly breathed in or lodged within the eye, whilst in the case of fomites, the virus infects through touching the face with contaminated hands [5]. Common *COVID*-19 symptoms include cough, fever, shortness of breath, and loss of smell [1]. In a limited number of cases, serious complications can develop, which include pneumonia and severe acute respiratory syndrome (*SARS*) [8], leading to a high mortality rate. Typically, symptoms appear around five days after the time of infection. However, it can take longer, and up to 80% of the patients may show no, or only mild, symptoms [8]. Although *COVID*-19 patients are most infectious within the first few days following the onset of the symptoms (post-symptomatic), both pre-symptomatic and asymptomatic patients, have been shown to be infectious, at least to some extent [9]. As no anti-*COVID*-19 treatment is currently available, non-pharmaceutical interventions based on isolation of both infected and non-infected people have thus far been the primary means for avoiding the spread of the virus [4, 10].

After several weeks of confinement, many countries are progressively restarting their economic and social activities. In order to avoid further spreading of the virus, the re-opening of the economy should ideally proceed in a fashion that minimizes interpersonal contacts. It is thus necessary to assess the risks of infection associated with different relaxation strategies [10-12]. In this paper, we introduce a stochastic coarse-grained model for the relaxation of lockdown that considers the re-opening of society in terms of people moving between sites on a one-dimensional grid. Our model contains three adjustable parameters characterizing viral transmission, viral detection, and degree of personal mobility. The results of the model will help in the qualitative assessment and semi-quantitative ranking of the importance of factors to be considered in the process of ‘restarting’ social and economic activities.

## Methods

To simulate a population under lockdown, we used a one-dimensional grid containing *M* sites into which *N* ‘people’ were distributed in either a regular periodic fashion or randomly without exclusion (Fig.1). During the lockdown period, each person’s mobility was restricted, and it was only allowed to move by a random distance (*m*) equal or less than the maximum range of mobility (*m*_max_). Thus, *m*_max_ = 0 represents a full lockdown whereas *m*_max_ = 1 describes the case where people can, with equal likelihood, either stay on the same site, move by one site to its left or to its right. Un-restricted mobility is attained when *m*_max_ ≥ (*M*−1)/2, (an odd number is used for *M* to avoid non-integer movements). Each movement was considered to take place in a time period *Δt*. The total time from the beginning of the lockdown was thus *t* = *Δt j*, where *j* is the number of periods completed up to time, *t*. A periodic boundary condition was applied to the running value of each *i*^th^ person’s position coordinate to suppress effects related to the finite size of the grid. Namely, (*x*_*i*_)_periodic_ = (*M* – |*x*_*i*_|) for *x*_*i*_ < 0; and (*x*_*i*_)_periodic_ = |*x*_*i*_ − *M*|for *x*_*i*_ > *M*. Simulations were run for *S* steps, and the results were averaged over *I* iterations. The following parameters were used to characterize viral transmission during the relaxation of lockdown.

**Figure 1a:**
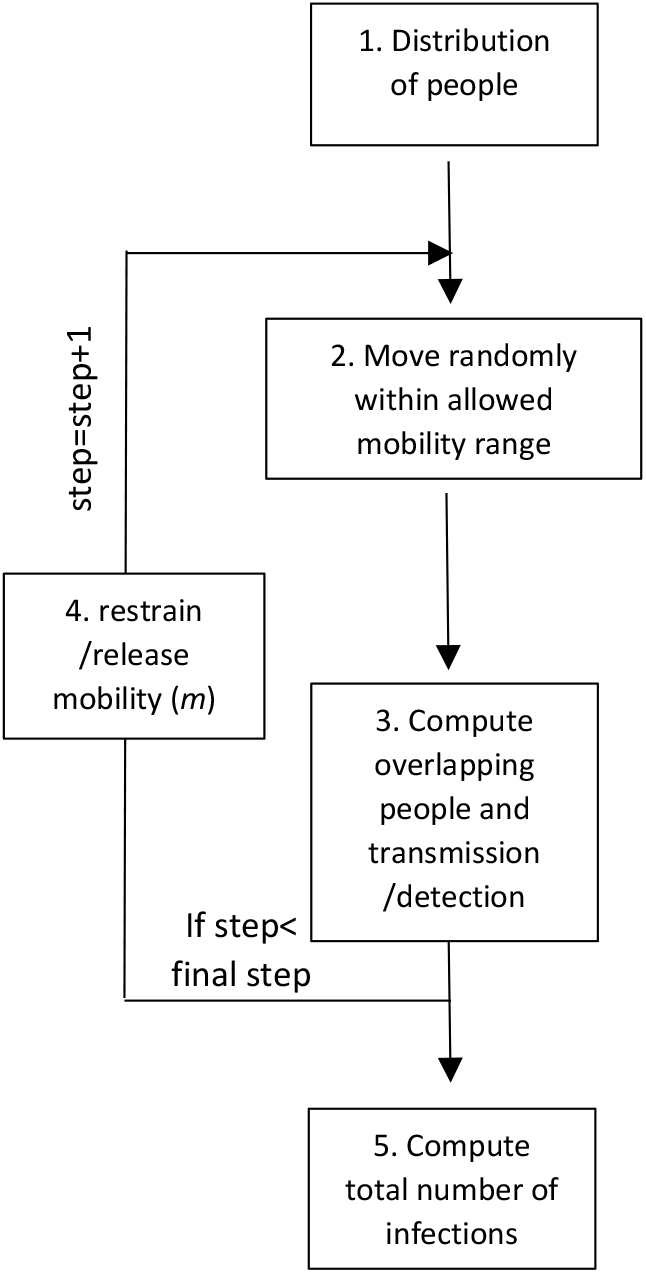
Schematic algorithm of the simulation. The main steps are as follows: 1. *M* (=1000) people are distributed on a one-dimensional grid having *N* sites (=2000). 2. They can move freely within a window of +/− *m* sites (*m* was set to 100 when no confinement was enforced). *m*=0 means that people were not allowed to move from their sites. 3. The people collocated at the same site were identified. People collocated with an infectious person were infected with a probability of *TP*, and the infected people were detected with a set detection probabililty (*DP*) and isolated from the system (detected people cannot infect nor be infected). 4. When the detected number of infected people reached 1% of the total number of all people (1% of 1000 people in our setting), *m* was decreased to the values indicated in Table 1. Infected people remained infectious for a total of 15 steps: 5 steps as a pre-symptomatic patient, and then ten steps as a symptomatic patient. Furthermore, we assumed that the virus would disappear after 15 steps, and that the infection occurs only once.

**Figure 1b:**
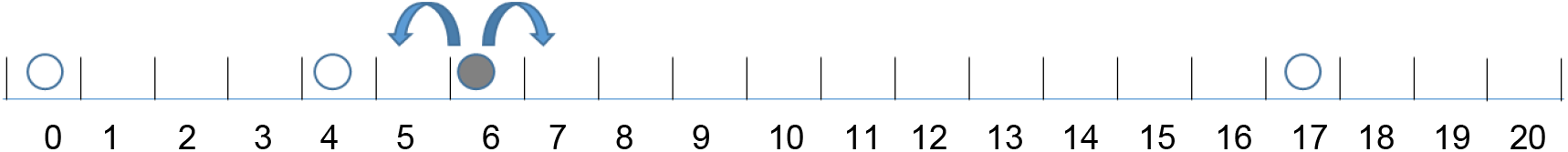
Schematics of the one-dimensional grid model with four people and 21 sites. The black and white circles represent the infected and the non-infected person, respectively. The arrows illustrate the possible movement of the infected person. For *m*=1, the person can move by −1, 0, or +1. Viral transmission occurs with a probability of *TP* when an infected person occupies the same sites as a non-infected person. The periodic boundary condition implies that a person at position 21 can move to its right and will “reappear” at position 0 (and vice et versa). It is used to alleviate the effects of the boundaries.

i. ***Transmission Probability (TP):*** *TP* describes the fractional probability of infection of a previously non-infected person by an infected person when the two are co-located simultaneously at the same grid site.
ii. ***Infection Period (IP):*** An infected person was considered to remain infectious for a given number of time intervals denoted by *IP*, after which the person was no longer considered infectious and considered to have acquired immunity (i.e., they can’t be infected a second time). Note that this study does not specify whether the patient recovers or not during, or at the end of the *IP*, as it is simply set apart the system (footnote 1).
iii. ***Pre-symptomatic Phase (PP):*** We assumed that following infection, *SARS-COV2* in infected people could not be detected (or detected with a lower sensitivity) for a certain time period, termed the pre-symptomatic phase (*PP*). In this report, we set *PP* to a period of 5*Δt* after the initial infection. Our model considers, in line with current medical observations, that infected patients can transmit the virus during this pre-symptomatic phase [10].
iv. ***Symptomatic Phase (SP):*** The period of the infection cycle from when the patient started to exhibit symptoms up until the illness was resolved, was termed the symptomatic phase. The *SP* period was considered to last for 10*Δt*, beginning after *PP* period of 5*Δt* and lasting until the end of the infection period (*IP*), so that the total period of infection (*IP*) is 15*Δt*.
v. ***Detection Probability (DP):*** During the *SP* period, the virus is discoverable with a set detection probability (*DP*) at each step. In the simulation, detected patients are immediately isolated (i.e., set aside from the system), after which they are not able to transmit the virus.
vi. ***Detection Probability during the pre-symptomatic phase (DP2):*** We considered that during the *PP* the virus is barely discoverable as it means that several asymptomatic people need to be tested. We thus introduced a second detection probability (*DP2*) for people in the *PP*. As for the symptomatic case, detected patients are immediately isolated and, therefore, unable to transmit the virus. Additional features, such as people violating confinement or the introduction of better and faster diagnosis tests, could be explicitly factored into the model using additional parameters, however, the current model can include, to some extent, such considerations through modulation of the existing parameters.
vii. ***Simulation Particulars*:** The programs were encoded using python **(3.4.8)**, and random numbers were generated using the python “rand” function. The programs were run on an 8 Xeon processor Linux server (HPC Systems, Tokyo, Japan). A 100 steps simulation with 1000 people and 2000 sites took between 90 and 190 seconds for completion, depending on the initial conditions.

## Results

### A. A probabilistic approach for simple settings (M=21 and N=3)

Simulation results were generated using a one-dimensional grid model. Analytical probabilities can be calculated for some simple limiting cases and we used them to check the veracity of the simulation (Supplemental materials). We first concentrated on clarifying the relationship between personal mobility following a period of lockdown (*m*_max_= 0) and the number of person-to-person contacts using a simplified setting (*M*=21 and *N*=3). We consider two general types of initial distribution (i) random, and (ii) regularly spaced.

i. Random initial distribution: As described in the supplementary section, when people are randomly distributed on the grid, the probability that two people are collocated at the same site at the beginning of the lockdown relaxation period (*t*=0) is given by *P*(2, *t*=0) and for *M*=21 and *N*=3 *it* is calculated as *P*(2,*t*=0)= 1/7 = 0.1428, (see Supplementary section A). Relaxation of personal mobility with random displacement will not disturb the initial random nature of the distribution, and so, on average, the encounter probabilities at a later time will remain unchanged (i.e., *P*(2, *t* >0) = 0.1428, see Supplementary section A). Note that this is an equilibrium average with deviations from the mean expected for any single simulation.
ii. Regularly-spaced initial distribution: We next consider how the encounter probability changes when the initial disposition is a regularly-spaced distribution whereby the three people are equally separated by seven distance units). For the regularly-spaced distribution, the probability of person-to-person contact at time zero is itself zero, i.e., *P*(2, *t*=0) = 0. After multiple steps with restricted or non-restricted random movement, we expect that the encountering probability will reach its random value of 0.1428 (for *M*=21, *N*=3), and this is indeed what we observe (Fig. 2). In order to investigate, in detail, the effects of mobility on the encountering probability, we calculated its value after a single time step, *P*(2,*t*=*Δt*) for mobility *m*_max_ ranging from 0(null) to 10 (full mobility). Due to their initial separation of six distance units, only mobilities (*m*) that can span this separation distance have the capability for producing a *P*(2, *t*=*Δt*) > 0, as seen in Fig. 2 for *m*_max_ ≥ 4 (Fig. 2 and Supplementary section C).

Another intuitive way of describing this point is to note the relationship between the probability for person-to-person overlap, *P*(2, *t*=*Δt*), and the person’s density, ρ_L_ = *N*/*M*, and personal mobility, *m*_max_, is given by (Eqn. 1).

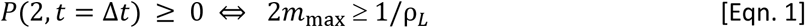

**Figure 2a:**
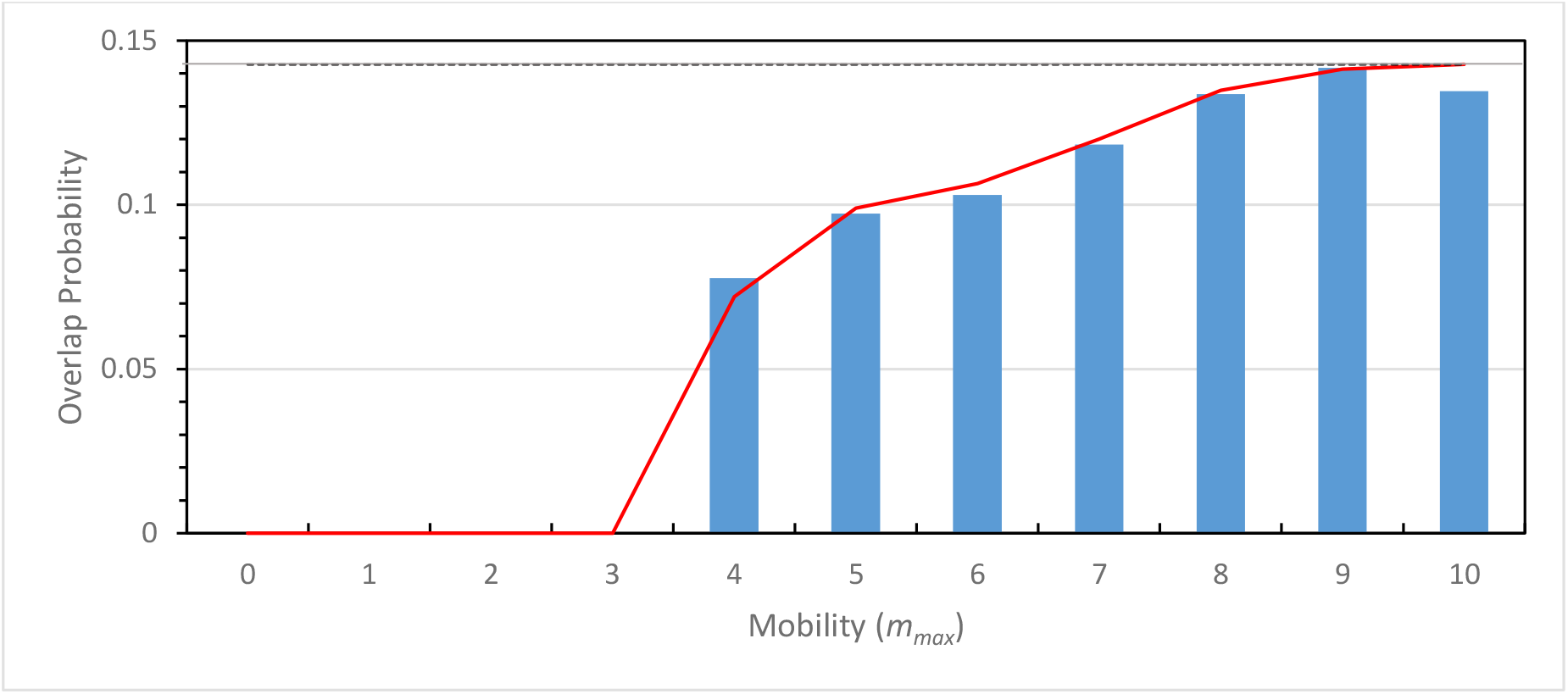
Comparison of the contact probability calculated using an exact probabilitic model (supplemental), and numerically from our simulation (average over 3000 runs with *N*=3, *M*=21). In the initial distribution, people were regularly-spaced at positions 0, 7, and 14 on the one-dimensional grid with a periodic boundary condition (between positions 0 and 20). People were then allowed to move according to any site elements within the mobility limit set to m_max_. The blue bars represent the results of the simulation, the red line those of the exact probabilistic mode, and the gray line represents the average probability of encounter for a random distribution (0.1428 for *N*=3 people on a grid with *M*=21 sites.

**Figure 2b:**
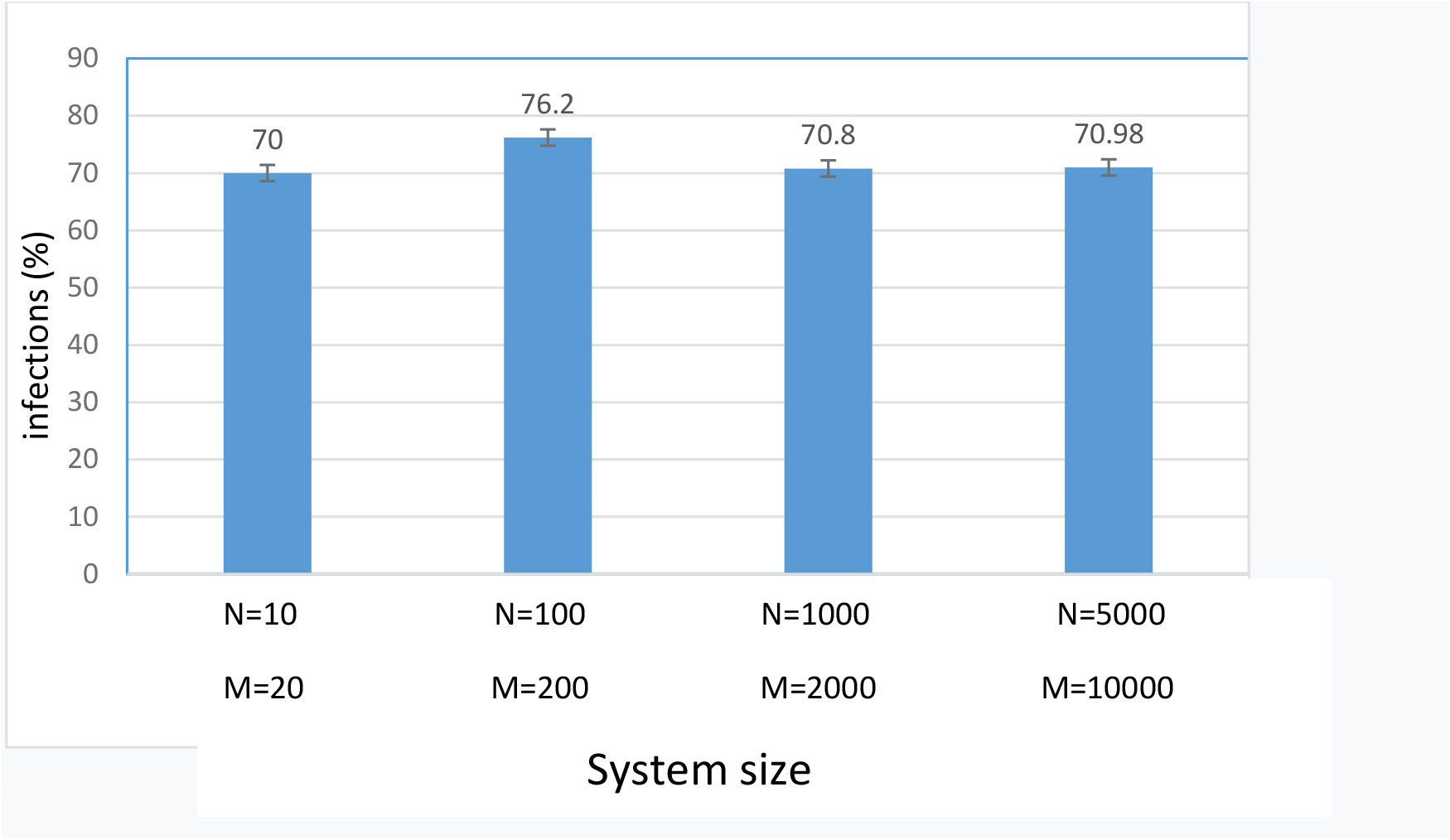
Size dependency. The percentage of infected patients was about 70% for systems with grid sizes ranging from 20 to 10000 positions, and a population density of 0.5. The values are averaged over 5 runs of 100 steps

Thus, for 2*m*_max_≥ 1/ρ_*L*_ the people can move freely, and it is thus expected that the contact number converges to the above asymptotic value of *P*(2,*t*=*Δt*)= 0.1428 (for *M*=21 and *N*=3)

### B. A one-dimensional grid model reproduces the basic features of the viral spread statistics

(*M*=2000 and *N*=1000)

Next, we examined whether our model can reproduce some basic features of the viral spread for a larger system. Thus, we first assessed that the results of our simulation are independent from the model’s size (or free from finite-size effects). To this end, we calculated the total number of infections for *M*= 20 to 10000. The number of infections were essentially constant at around 70% of the total number of people, indicating the validity of the simulation within the framework of the model (Fig. 2b).

Using a model with 1000 people and a grid of 2000 positions, we then showed that our model reproduces the basic features of an infection outbreak starting from a single pre-symptomatic individual seeded randomly into the system (Figure 3). In the beginning, mobility was not restricted, and people could move within a mobility range set to *m*_max_ = 100. In our model, the lockdown was simulated by reducing the mobility from the initial *m*_max_ = 100 to 0 (or to the value given in Table 1), when the number of detected infections reached 1% of the population (10 people). This mobility restriction was relaxed after an interval of 10*Δt*, which corresponds to the time of symptomatic infection in our model. As can be noted, during the early stages, the number of infections remains low (until step 10 in Figure 3), but after *t* = 20*Δt*, the number of new patients increases significantly (despite the average number of contacts remaining the same). An apparent second wave appears due to the fact that many undetected infectious patients remain within the system after the end of the first wave. It is noteworthy that at the end of the simulation, some people remain un-infected.

**Table 1a:** Dependency of the number of infections on mobility restriction, detection probability, and transmission probability (*N*=1000 People, *M*=2000 sites, 80% transmission probability (*TP*), mobility restriction applied at 1% (=10 people))

| **Detection Probability** | **Tot Infect <sup>1</sup>** | **Tot Infect <sup>1</sup>** | **Tot Infect <sup>1</sup>** | **Tot Infect <sup>1</sup>** |
| --- | --- | --- | --- | --- |
| **0%** | 998 | 997 | 998 | 997 |
| **20%** | 613 | 879 | 909 | 946 |
| **40%** | 189 | 727 | 834 | 906 |
| **60%** | 71 | 601 | 698 | 847 |
| **80%** | 51 | 521 | 567 | 810 |
| **100%** | 49 | 341 | 426 | 797 |

**Table 1c:** Dependency of the number of infections on mobility restriction and detection probability, a d transmission probability (1000 persons, 2000 sites, 60% detection probability, restriction at 1%)

|  | **Total Infection <sup>1</sup>** |  |  |  |
| --- | --- | --- | --- | --- |
| **Transmission Probability** | **$m_{\max}=0$** | **$m_{\max}=3$** | **$m_{\max}=5$** | **$m_{\max}=100$**<br>**(initial)** |
| **40%** | 32 | 64 | 127 | 305 |
| **60%** | 59 | 247 | 580 | 710 |
| **80%** | 81 | 569 | 667 | 837 |
| **100%** | 87 | 673 | 797 | 919 |

1. Total number of particles infected at the end of the simulation.

**Table 1c:** Dependency of the number of infections on mobility restrictionand detection probability during the presymptotic phase, and transmission probability (1000 people, 2000 sites, 80% transmission probability, restriction at 1%) Total number of infection for DR2= 0 to 40%

**Total number of infection for DR2= 0 to 40%**
| | $m_{\max}=0$, Tot inf<sup>2</sup> | | | $m_{\max}=3$, Tot inf<sup>2</sup> | | | $m_{\max}=5$, Tot inf<sup>2</sup> | | | $m_{\max}=100$, Tot inf<sup>2</sup> | | |
| --- | --- | --- | --- | --- | --- | --- | --- | --- | --- | --- | --- | --- |
| Detec Prob<br>$PP^1(DP2)$ | Tot<br>$DR=80\%$ | Tot<br>$DR=60\%$ | Tot<br>$DR=40\%$ | Tot<br>$DR=80\%$ | Tot<br>$DR=60\%$ | Tot<br>$DR=40\%$ | Tot<br>$DR=80\%$ | Tot<br>$DR=60\%$ | Tot<br>$DR=40\%$ | Tot<br>$DR=80\%$ | Tot<br>$DR=60\%$ | Tot<br>$DR=40\%$ |
| **0%** | 59 | 66 | 110 | 323 | 562 | 669 | 452 | 574 | 792 | 815 | 863 | 897 |
| **5%** | 38 | 47 | 57 | 201 | 280 | 578 | 273 | 299 | 632 | 713 | 739 | 841 |
| **10%** | 23 | 26 | 32 | 110 | 128 | 202 | 111 | 124 | 381 | 599 | 653 | 757 |
| **20%** | 13 | 21 | 28 | 23 | 33 | 44 | 45 | 67 | 76 | 196 | 223 | 211 |

**1.** DR2: Detection probabilities at the presymptotic phase. **2.** Total number of particles infected at the end of the simulation.

**Figure 3.**
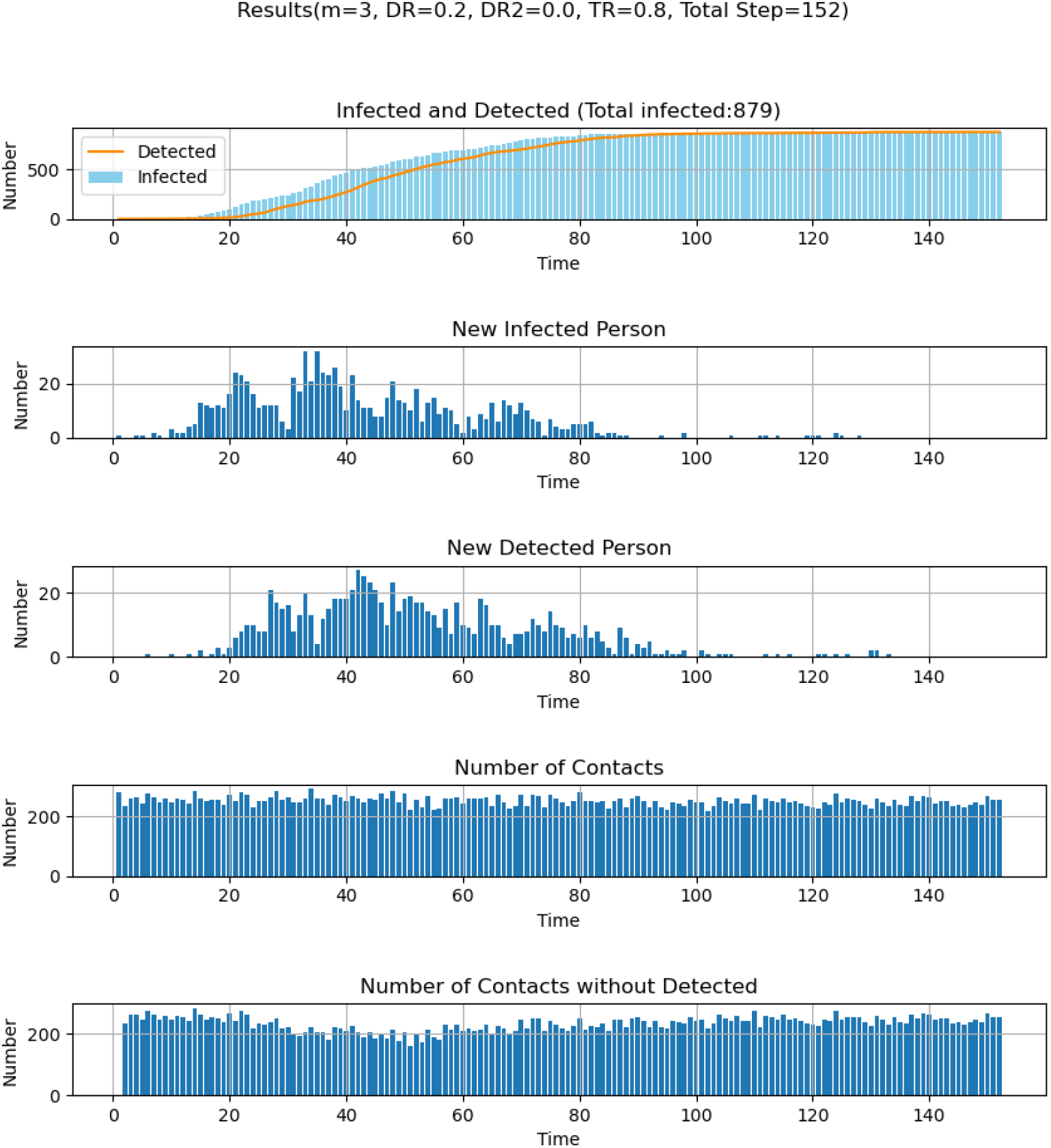
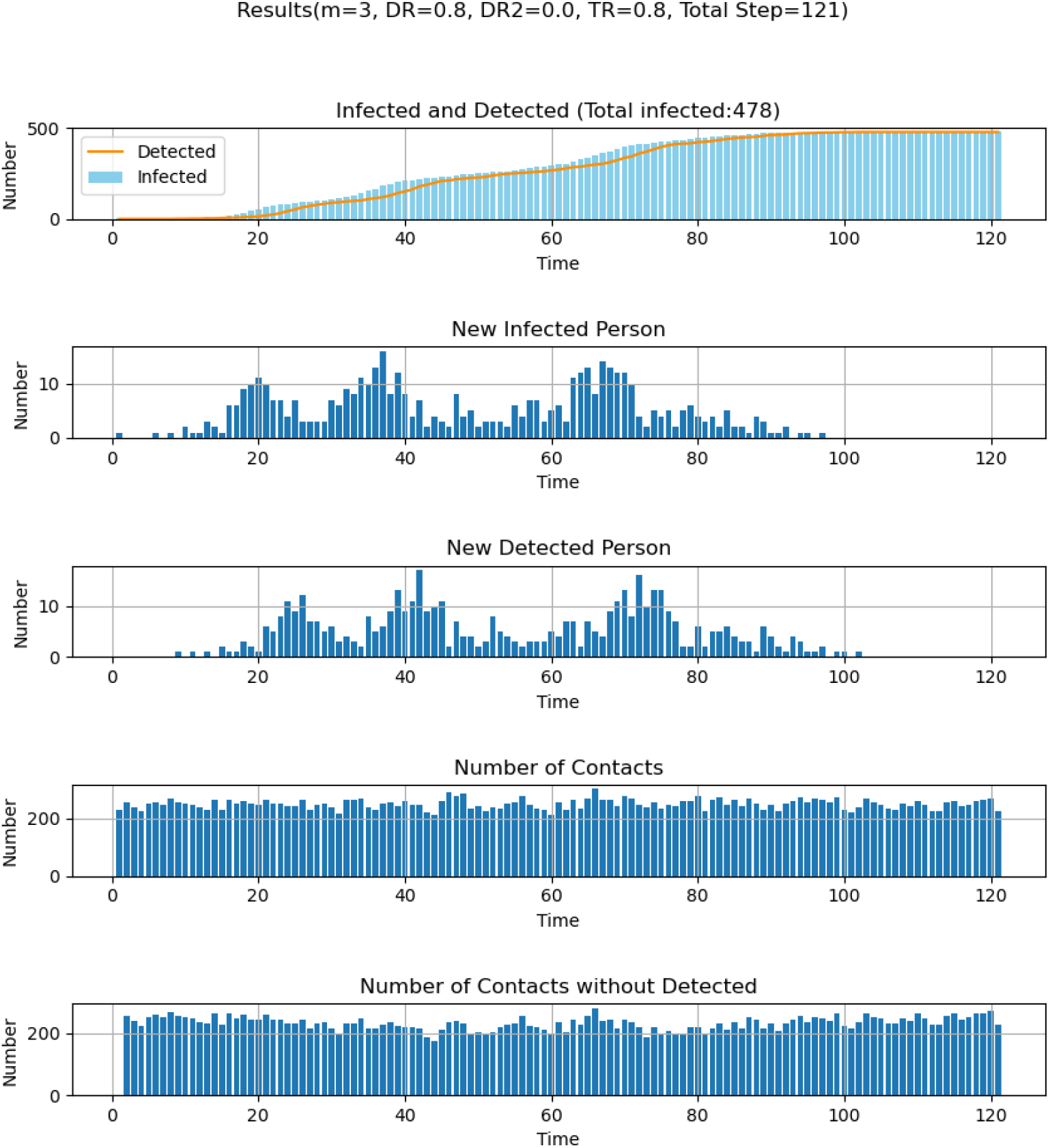
**(A)** Examples of time-dependent virus transmission. The parameters are as follows: *N*= 1000, *M*=2000, t=1. At the initial state, *m*_max_ was set to 100 and is reduced to *m*_max_ = 1 when the number of detected infections reaches 1% (10 people). Infected patients are infectious for 15 steps (five steps as non-detectable asymptotic persons and ten steps as a symptomatic and detectable patient). After fifteen steps, they are removed from the system: they cannot be infected nor become infectious a second time. Parameters are given in the figures.

This is because, in our current setting, we assumed that people are infected only once, and remain infectious for only fifteen time-steps. Under this setting, the spread of infection stopped on average after *t* = 70*Δt* because no infectious patients remained in the system (Figure 3).

### C. Effect of mobility restriction vs. reduction of viral transmission and the probability of detection

Next, we compared the effect of change in mobility, *m*_max_, against variation in either of the two parameters relating to detection probability (*DP and DP2*) and transmission probability (*TP*) (Table 1). Similar to the simulations shown in section B, a partial lockdown involving a change in *m*_max_ was instituted when the number of detected infections reached 1% of the population.

To investigate the importance of mobility vs. detection probability, we performed simulations, in which the detection probability for symptomatic persons (*DP*) was varied from 0 to 1 for five mobility values. In this simulation, the detection probability for pre-symptomatic cases (*DP2*) was set to zero. *DP* is interpreted as the probability that a symptomatic patient is correctly diagnosed and subsequently isolated (meaning they cannot then infect other people). A null detection probability (*DP* = 0) means that symptomatic patients are neither diagnosed nor isolated. On the other hand, a 100% successful detection protocol (*DP* = 1) implies that all suspected symptomatic patients are systematically tested and diagnosed with perfect accuracy; and that once identified, the patients are strictly isolated. Our results suggest that the reduction of mobility is a determining factor, but that full lockdown (*m*_max_=0) has to be coupled with a detection probability > 0.4 to be effective (Figure 4, Table 1b). When the detection probability falls to 0.2 or less, the beneficial effects of the lockdown are significantly reduced (Figure 4, see *m* = 0) with the total number of infections eventually increasing to values comparable to a scenario involving no lockdown but a detection probability of 0.6 to 0.8 (Fig 4, *m*_max_=5). Note that the high infection levels observed for *DP* = 0 are somewhat misleading because, in our simulation, the infected patients are not detected nor isolated (*DP* = 0), and therefore the lockdown value of 1% of the population is never reached, and thus no lockdown implemented during the entire simulation.

**Figure 4.**
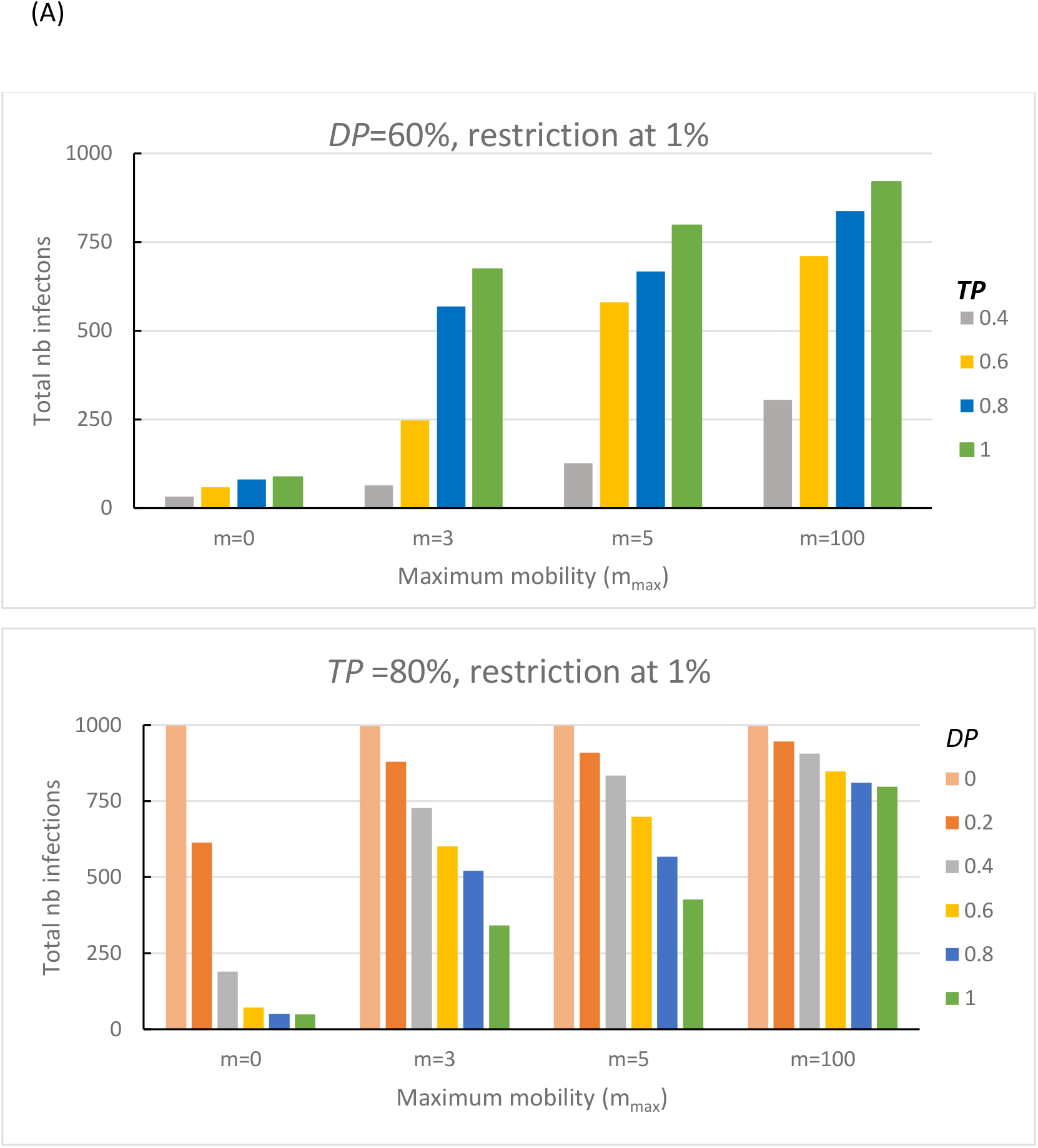
Effects of mobility restriction vs. Detection Probability (*DP*) and Transmission Probability (*TP*) on infection probability. The other parameters are as follows: *N*= 1000, *M*=2000 and during the non-confinement period, the mobility (m_max_) is set to 100. The mobility was constrained when the number of detected infections reaches 1% (10 particles) by setting max (*m*)) to the indicated value (*m*). People that are not detected during the 15 days of infection (5 days of asymptotic and 10 days of symptomatic infections) won’t be infectious nor be infected and are practically set apart from the system (A) *DP* was varied from 0.0 to 1.0 with a *TP* set to 0.8. (B) *TP* was varied from 0.4 to 1.0 using a *DP* = 0.6. Color codes for *DP* and *TP* are given within the figures.

To investigate the importance of mobility vs. transmission probability, we performed simulations in which the transmission probability (*TP*) was varied from 0.4 to 1 for five mobility values. The transmission probability (*TP*) is the fractional probability of infection of a non-infected person by an infected patient when the two are co-located at the same grid site during a single time period. In real life, the *TP* parameter can be controlled by non-pharmaceutical measures [13, 14] such as social distancing[15-17], hand washing, and mask-wearing [18, 19]. As expected, our simulations show that reducing the transmission probability has a considerable role in decreasing the total number of infections (Figure 4b). Less expected was the finding that reducing *TP* to a value of 0.4 in combination with enforcing reduced mobility measures was as effective as enforcing a complete lockdown (Figure 4b; *m*=0). Inversely, a high *TP* (≥ 0.6) strongly increased the spread of the virus (Figure 4b).

Finally, we examined the effect of detecting the virus at the pre-symptomatic phase (***PP)*** by setting *DP2* > 0. This approach requires that a high fraction of the entire population to be tested on a daily basis, which might not be sustainable on a large scale. However, this strategy proved extremely effective because infected patients in the *PP* period were detected and put in quarantine very early in the infection cycle. Additionally, since the mobility reduction is decided based on the number of detected infections (rather than the actual number of infected patients), identifying these pre-symptomatic patients early on can more assist in more closely estimating the actual number of infections in the system, thus implementing an earlier lockdown. Noteworthy, increasing *DP2* from 0 to just 10% can dramatically decrease the number of infections, even for a mild mobility restriction of *m*_max_ = 3 or 5 (Table 1c).

## Discussion

The spread of viral infection is usually modeled using analytical models developed with different levels of complexity [12]. In contrast, the one-dimensional numerical approach introduced here has the merit of simplicity, and as such it enables to readily comprehend our parameters to real-life features; and it is able to offer both qualitative and semi-quantitative predictions for more sophisticated scenarios. For example, we could readily accommodate a population with two types of people exhibiting mobility and transmission characteristics representative of, for instance, the young and the older generation. Similarly, the model can be readily adapted to fit other bespoke confinement /de-confinement strategies for particular social situations. Such versatility is frequently unattainable with an analytical model, which tends to be generally less flexible [11, 12, 20]. Another advantage of using numerical simulation is that parameters, such as the reproduction ratio [20, 21], are straightforwardly determined via exercises in counting.

As with all simulations, the interpretation of the results requires caution. Seeking exact correspondences in the real world for our generalized descriptive variables and parameters (e.g., those relating to unit time (*Δt*), physical collocation, and mobility) involves a certain degree of ambiguity. Nevertheless, despite these limitations, we believe that our model provides useful qualitative and semi-quantitative information on how virus-specific and society specific parameters influence the way viral spread occurs during periods of lockdown and re-opening. While some observations were anticipated, such as an early lockdown being more effective than a delayed one, others were less intuitive. For example, the relationship between mobility and the probability of encounters were indeed critically dependent upon both the initial distribution of the population and the population density. Furthermore, our model predicts that the mobility restriction must be stringently enforced and accompanied by a high detection/isolation probability in order to reduce the total number of infections significantly. Finally, the detection and quarantine of pre-symptomatic patients (which would require testing a large number of people every day and is therefore not a realistic strategy) would reduce the final number of infections by a factor of ten or more. Successful strategies for achieving a low number of infections include full confinement combined with a reasonable detection probability of symptomatic patients (*DP* > 0.4). A realistic strategy might be mild confinement, with a high detection probability during the symptomatic phase and a reasonable detection probability during the pre-symptomatic phase (for example, *m* =5, *DP* =0.8, *DP2* = 20 in Table 1c. Such strategy is in line with a recent report by Muller et al.[22]). Variation in such factors may be the reason behind the wide variation of infection numbers observed in different countries [23].

## Conclusion

We presented a stochastic coarse grained model where people were allowed to either move freely or in a constrained manner, and viral transmission can occur when infected and non-infected individuals overlap at the same site. Our model includes adjustable parameters characterizing viral transmission probability, detection probability, and personal mobility within a population. Importantly, our simulation can reproduce the basic aspects of viral spread within a community and the development of a local epidemic. The results of our simulation agreed well with exact probabilistic calculations, which, under certain simple limiting cases, were readily formulated. Although many of the results were in line with our anticipation, our model revealed a number of interesting features. In particular, we noticed that the link between personal mobility and the risk of encounter (and thus infection) is a step function when the initial distribution is regularly-spaced, and the maximum mobility lies below the inverse spatial density of the population. Below this critical juncture, the risks of infection were zero, whereas at greater mobility (or higher population density) the situation rapidly approached the random case. The approach described here provides a qualitative assessment of the efficacy of modifying societal parameters that should prove useful to decision-makers when considering the relaxation of lockdown conditions.

## Data Availability

All data are provided in the supplemntal data

## Acknowledgments

This research was motivated by an intense discussion with Prof. Christopher Mudry (Paul Scherrer Institute and Ecole Polytechnique de Lausanne, Switzerland), for which Y.K. is very thankful. We thank Ms. Jingwen Xian and Mr. Zhirui Cheng for discussion and Python programming of the early versions. D.H. would like to thank the Nagoya Institute of Technology for a placement on their Visiting International Scientist Program.

## Funding

This research was supported by a JSPS grant-in-aid for scientific research (KAKENHI, 15H04359, and 18H02385)

## Competing interests

The authors declare no competing interests.

## Data Sharing

All data are given in the manuscript and the supplementary data. The original program can be freely accessed at http://domserv.lab.tuat.ac.jp/covid19.html (under preparation)

## Authors Contribution

Y.K., and Y.M. designed the project, Y.K., Y.M., and D.H. derived the equations described in the appendix, Y.K., and D.H. wrote the manuscript, H.T. and T.M. discussed the results and advised with paper writing, Y.M. and S.A. wrote the program performed simulation. All authors read and approved the manuscript.

## Footnote1

We are aware of a number of reports describing the potential for *COVID*-19 reinfection of previously recovered patients [[24-26]]. Although this situation is considered uncommon, the simulation could be readily modified to include such a scenario.

## Notes

### Competing Interest Statement

The authors have declared no competing interest.

### Funding Statement

JSPS (Japanese Society for the promotion of Science)

